# A Network Analysis Framework to Improve the Delivery of Mosquito Abatement Services in Machala, Ecuador

**DOI:** 10.1101/19009498

**Authors:** Catherine A. Lippi, Liang Mao, Anna M. Stewart-Ibarra, Naveed Heydari, Efraín Beltrán Ayala, Nathan D. Burkett-Cadena, Jason K. Blackburn, Sadie J. Ryan

## Abstract

**Background:** Vector-borne disease places a high health and economic burden in the American tropics. Comprehensive vector control programs remain the primary method of containing local outbreaks. With limited resources, many vector control operations struggle to serve all affected communities within their districts. In the coastal city of Machala, Ecuador, vector control services, such as application of larvicides and truck-mounted fogging, are delivered through two deployment facilities managed by the Ecuadorian Ministry of Health. Public health professionals in Machala face several logistical issues when delivering mosquito abatement services, namely applying limited resources in ways that will most effectively suppress vectors of malaria, dengue, and encephalitis viruses.

**Methods:** Using a transportation network analysis framework, we built models of service areas and optimized delivery routes based on distance costs associated with accessing neighborhoods throughout the city. Optimized routes were used to estimate the relative cost of accessing neighborhoods for mosquito control services in Machala, creating a visual tool to guide decision makers and maximize mosquito control program efficiency. Location-allocation analyses were performed to evaluate efficiency gains of moving service deployment to other available locations with respect to distance to service hub, neighborhood population, dengue incidence, and housing condition.

**Results:** Using this framework, we identified different locations for targeting mosquito control efforts, dependent upon management goals and specified risk factors of interest, including human population, housing condition, and reported dengue incidence. Our models indicate that neighborhoods on the periphery of Machala with the poorest housing conditions are the most costly to access. Optimal locations of facilities for deployment of control services change depending on pre-determined management priorities, increasing the population served via inexpensive routes up to 34.9%, and reducing overall cost of accessing neighborhoods up to 12.7%.

**Conclusions:** Our transportation network models indicate that current locations of mosquito control facilities in Machala are not ideal for minimizing driving distances or maximizing populations served. Services may be optimized by moving vector control operations to other existing public health facilities in Machala. This work represents a first step in creating a spatial tool for planning and critically evaluating the systematic delivery of mosquito control services in Machala and elsewhere.

## Background

### Public Health Vector Control in Latin America

Delivery of vector control services in the public health sector is challenging throughout much of Latin America, where the management of vectored diseases is complicated by diversity in both pathogens and vectors, most notably in the tropics. High prevalence of mosquito-borne diseases, coupled with oftentimes limited capacity for mosquito abatement and medical services, can rapidly overwhelm existing healthcare systems [1–4]. Still, outbreaks of mosquito-borne pathogens are problematic, even in municipalities with excellent public health infrastructure. Comprehensive vector control programs are widely acknowledged as a fiscally conservative strategy for suppressing and preventing outbreaks of mosquito-borne diseases in Latin America, employing combinations of surveillance, abatement, and educational outreach to the public [5–9]. Indeed, the operating budgets of vector control agencies pale in comparison with the resources consumed, and productivity lost, during large outbreak events. Nevertheless, the costs incurred by vector control efforts can still pose significant burden, particularly in communities with limited funds dedicated to public health activities [10].

The consequences associated with implementing control strategies without robust planning and review are many, ranging from reduced impact of funding streams to outright intervention failure [11,12]. It is therefore imperative that vector control agencies critically plan and evaluate their delivery systems to ensure efficient operations and judicious application of resources. There are two strategies of applying public health vector control efforts to control outbreaks: i) proactively, where high risk areas of mosquito production are targeted based on prior information ahead of transmission peaks, and ii) reactively, where abatement activities are triggered in direct response to incoming surveillance data, particularly high numbers of human disease cases [13]. Although well-planned proactive vector control can be immensely advantageous, effectively reducing mosquito source populations and suppressing transmission before outbreak events occur, public funding is often skewed towards reactive programs, where the rapid deployment of service is triggered by reported disease clusters [13].

### Vector Control and Mosquito-Borne Diseases in Ecuador

Located on the northwestern coast of South America, Ecuador has historically been an active area of mosquito-borne disease transmission, long contending with seasonal outbreaks of malaria and dengue. Much of this seasonal transmission is concentrated in densely populated areas of low elevation along the coast. Ecuador has a strong precedence of vector control activities, having formerly eradicated *Aedes aegypti*, and as a result yellow fever and dengue, in the 1950s [14]. However, eradication was followed by a period of lax vector control policy and diverted funding throughout Ecuador and much of South America, culminating in large outbreaks of dengue fever beginning in the late 1980s [3,15]. Currently, there is active transmission of several arboviruses in Ecuador, including yellow fever virus (YFV), four serotypes of dengue virus (DENV 1-4), chikungunya virus (CHKV), and Zika virus (ZIKV), all of which are competently vectored by the yellow fever mosquito (*Aedes aegypti*) [16–18]. The recent documentation of the Asian tiger mosquito (*Aedes albopictus*) in Guayaquil, Ecuador raises further concern with local public health officials, as this species is also capable of vectoring the same viruses as *Ae. aegypti* in Ecuador [19,20]. Despite the presence of competent vectors, targeted control has the potential to mitigate the effects of disease outbreaks, as was seen with the local elimination of malaria transmission at the Ecuador-Peru border, thus demonstrating the efficacy of consistently applied case surveillance and vector control programs [21].

Machala is a port city located in Ecuador’s El Oro province on the southern coast (Fig. 1). With a projected population of over 280,000, it is the fourth largest city in the country, second largest port, and a center of agricultural trade [22,23].

**Fig. 1.**
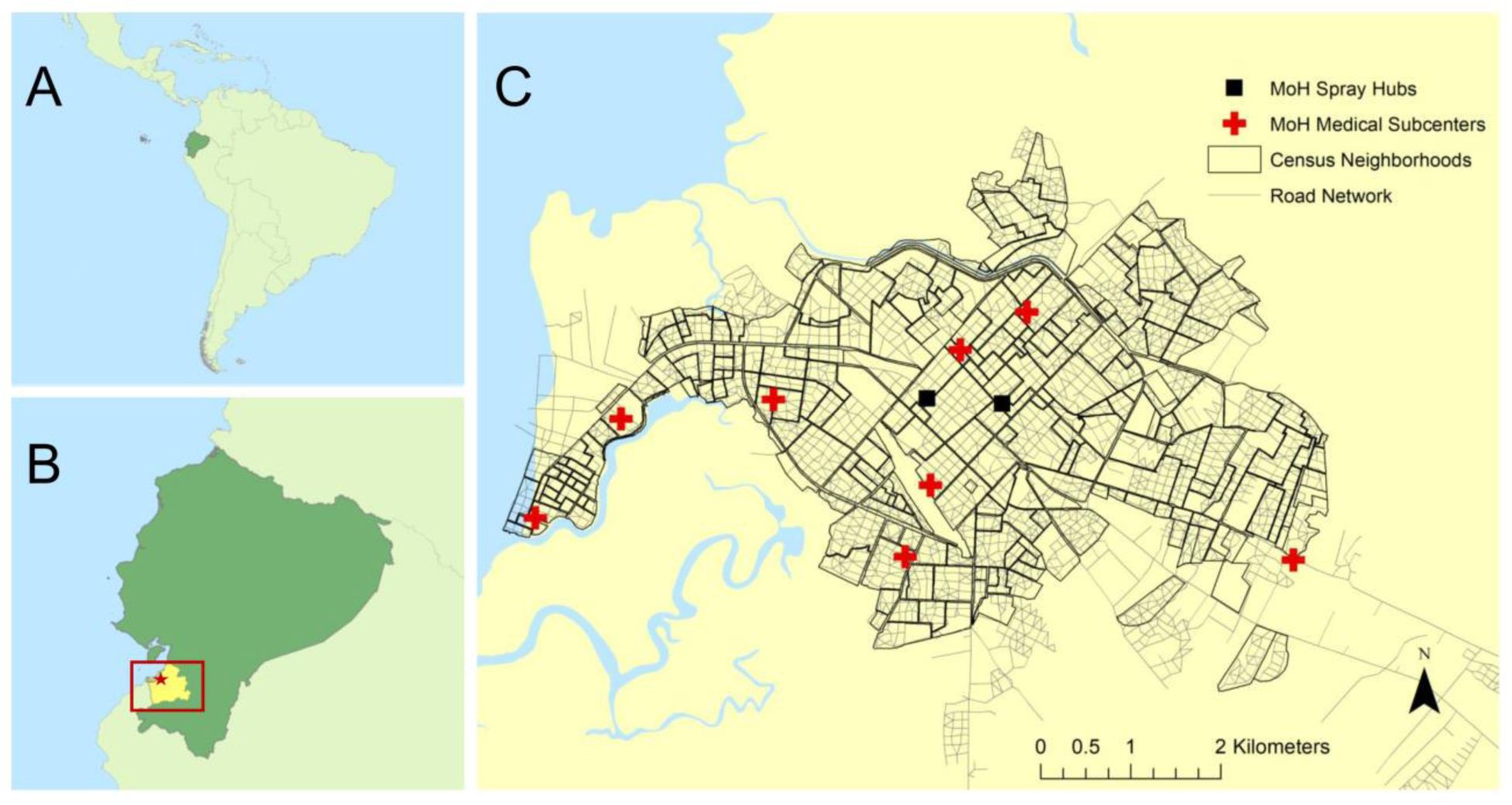
The South American country of Ecuador (A) contends with endemic dengue transmission, particularly in southern coastal El Oro province (B, shown in yellow). Machala (B, red star) is a port city in El Oro and the fourth largest city in the country. The Ecuadorian Ministry of Health deploys mosquito control activities in Machala through two centrally located deployment hubs (C). Mosquito abatement services formerly operated from several medical subcenters (red crosses) throughout the city. This figure was produced in ArcMap 10.4 (ESRI, Redlands, CA) using shapefiles from the GADM database of Global Administrative Areas, ver. 2.8 (gadm.org), transportation network and census data from INEC, and georeferenced facility locations provided by the MoH.

Machala has a long history of operational mosquito control and surveillance due to high dengue incidence relative to surrounding areas, making it an ideal setting to study delivery systems of vector control services. Formerly, fumigation and control services were deployed from decentralized medical subcenters located throughout Machala as a part of the National Service for the Control of Vector-borne Diseases (in Spanish: Servicio Nacional de Control de Enfermedades Transmitidas por Vectores Artrópodos – SNEM), a vertically structured vector-borne disease eradication program in the Ministry of Health (MoH) [24]. In 2015, SNEM was disbanded and public health surveillance and mosquito control in Machala became decentralized and administered by the local MoH health districts, with two centrally located hubs from which abatement services are deployed (Fig. 1). A variety of chemical control methods are utilized by these service hubs including aquatic larvicides (temefos/abate) and indoor residual spraying of residential homes (deltamethrin) delivered by technicians visiting homes, and ultra-low-volume (ULV) fogging with adulticides (malathion) delivered at the street level via trucks.

Implementation of vector control services in Machala requires considerable daily transportation, moving people, materials, and equipment from service hubs throughout the city via MoH trucks. Previous studies on dengue in Machala have shown associations between census housing characteristics and disease incidence, indicating that risk of contracting mosquito-borne diseases is not evenly distributed throughout the city [16,25]. Under current management strategies, neighborhoods are not prioritized for treatment in a proactive, systematic manner based on household-level risk factors, due in part to limited funding and resources [10]. Instead, service schedules are developed from local knowledge and experiences, with the goal of ensuring coverage to as many households as possible before and during the rainy season. Additionally, during transmission season spray treatments are delivered when surveillance cases exceed predetermined thresholds, or in response to residential mosquito complaints. While this method of service delivery is targeted in a sense, it is nevertheless in reaction to detected caseloads and self-reported mosquito presence, creating a lag between transmission and vector control, and potentially failing to treat high-risk neighborhoods with low reporting. Ideally, mosquito control operators in Machala should have tools available to plan control efforts in a more systematic and dynamic manner, emphasizing delivery of services to areas within the city with the highest risk of experiencing outbreaks.

### Transportation Network Analysis

Network analysis frameworks have long been used in the realm of public health planning to effectively allocate resources, improve operations, and guide policymaking in communities [26–28]. This family of analyses is particularly useful in the assessment of service demand, planning of delivery routes, and evaluation of deployment facilities in relation to underlying road networks. Many transportation network problems are based on road network distances, for example, establishing areas of service based on driving distance and finding optimal driving routes with a modification of Dijkstra’s algorithm, wherein the shortest distance paths between a given origin and destination pair are found [29,30]. Under this framework, the relative costs and potential benefits of service deliveries can be weighed under various management goals and priorities, providing a flexible tool to aid in proactive decision-making and resource allocation.

Although commonly used in the context of solving accessibility and allocation problems in the public health sector, to our knowledge there have been no efforts to apply network analysis methodologies in the optimization of vector control service delivery. Our goal was to build a network-based analytic framework that would aid in the planning and delivery of mosquito control services in Machala, demonstrating the utility of network analysis in a public health vector control context. With this goal in mind, the objectives of this study were to 1) establish service areas of vector control service deployment facilities based on road-network distance, describing factors that guide management decisions in context of accessibility; 2) identify the optimal delivery routes from current spray facilities to neighborhoods, estimating the relative costs of delivery; and 3) explore alternate locations of service deployment hubs under different management priorities, wherein we represent scenarios of proactive and reactive abatement schemes.

## Methods

### Data Sources

Census data collected in Machala, Ecuador, aggregated to neighborhood census blocks (n=254, referred to as neighborhood hereinafter), were provided by the Ecuadorian National Institute of Statistics and Census (Instituto Nacional de Estadística y Censos – INEC) for the most recent national census, conducted in 2010 [22,31]. Based on previous studies of dengue risk in Machala, factors of interest deemed relevant in relation to the delivery and prioritization of mosquito control services included data on population (Fig. 2A) and housing condition index (HCI) (Fig. 2B) [25]. Housing condition has been shown to be strongly associated with mosquito-borne disease incidence in Machala, as houses in poor condition allow mosquitoes to enter the home and have more abundant larval habitat in the home and patio [25,32,33]. The HCI is an aggregate variable combining grades of roof, wall, and floor quality as a measure of overall housing condition, with 0 being excellent condition and 1 being very poor condition.

**Fig. 2.**
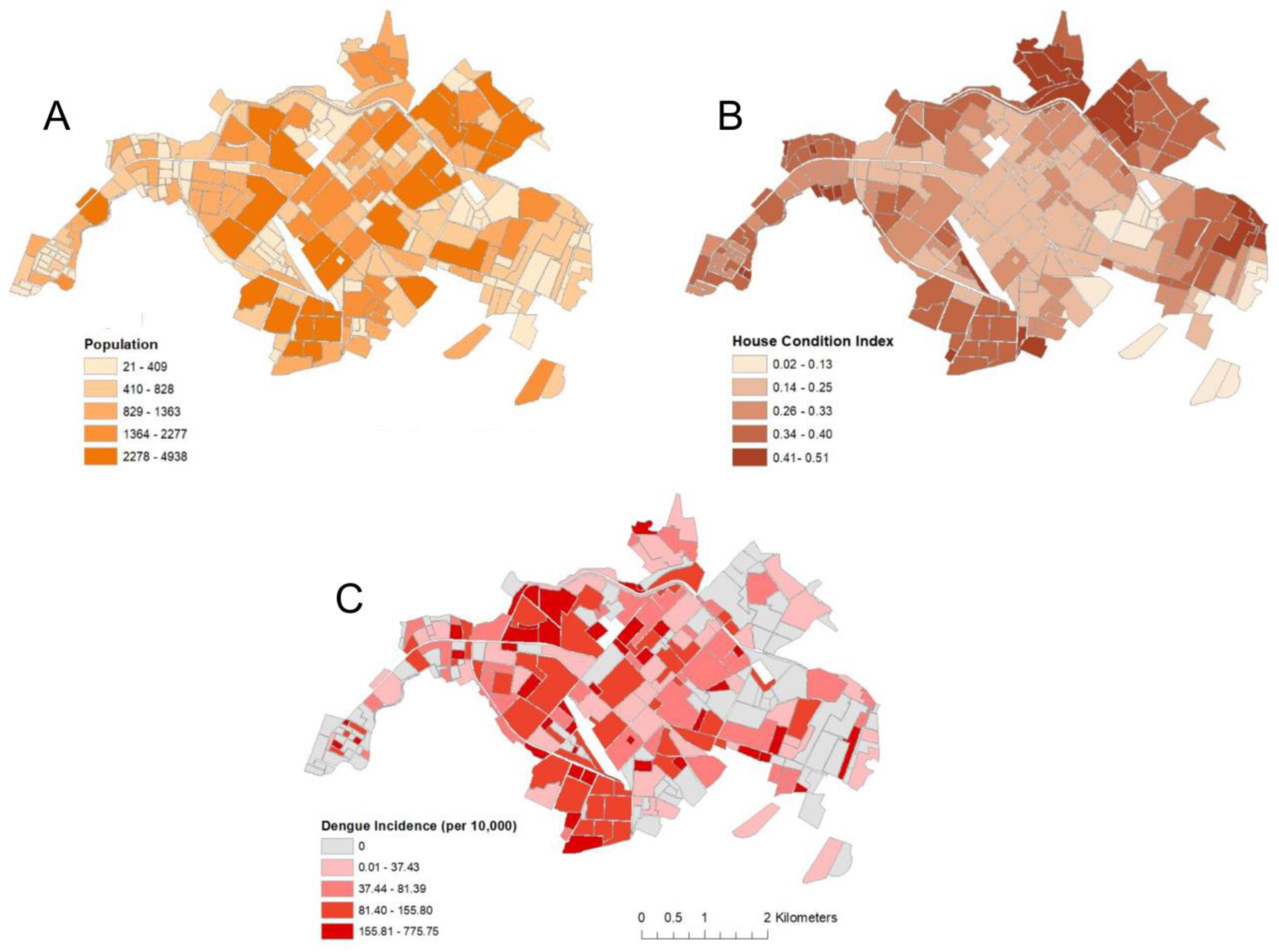
Census variables (INEC 2010) aggregated to the census-block level in Machala, Ecuador including (A) population and (B) housing condition index (HCI). The Ecuadorian Ministry of Health provided data on (C) dengue incidence in Machala for the year 2010. This figure was produced in ArcMap 10.4 (ESRI, Redlands, CA).

Data regarding the road network in Machala were also obtained from INEC, reflecting the most complete dataset for transportation networks available for the city. Although some information on road classification (e.g. primary versus secondary roads) was available, the majority of road segments are not classified. Consequently, all roads were considered to have the same functional accessibility in network analyses. Furthermore, Ecuador enforces uniform speed limits (50 km per hour) for all municipalities throughout the country, thus precluding the calculation of variable travel times [34].

De-identified human case data from a 2010 outbreak of dengue fever in Machala, aggregated to barrios (i.e. neighborhood census blocks), were provided by the Ecuadorian Ministry of Health (MoH) and have been previously described [25]. Human surveillance data are routinely used in making reactionary vector control decisions, and were used in our analyses as a weighting factor to simulate prioritization of service delivery. The MoH provided GPS coordinates for their two active mosquito abatement facilities, from which mosquito control services are deployed, as well as coordinates for eight MoH-operated medical clinics found throughout Machala, from which mosquito control services were formerly delivered. Census and human case data were mapped in ArcGIS (ver. 10.4) to visualize spatial patterns (Fig. 2C), serving as points of comparison and weighting factors for network analyses.

### Network analysis framework

*Establishment of Service Areas –* Spatial analyses of the transportation network in Machala were performed in ArcGIS (ver. 10.4) using the ‘Network Analyst’ extension toolbox. Service areas were generated based on driving distance from the two mosquito control facilities along the road network, enabling the identification of characteristics of the population served. Overlap of service area boundaries between the two spray hubs was allowed, and service areas were delimited at 0.5, 1, 3, 5, and >5 km driving distances from either facility. The census data were overlaid onto service areas to reveal population, housing characteristics, and reported dengue incidence (Table 1).

**Table 1.** Distribution of population, mean HCI, and 2010 dengue incidence by service area ranges.

| Service Area Range<br>(km) | Neighborhoods | Population | Mean HCI | Dengue Incidence<br>(per 10,000) |
| --- | --- | --- | --- | --- |
| **0.0 – 0.5** | 9 | 10,410 | 0.21 | **102.85** |
| **0.6 – 1.0** | 17 | 25,372 | 0.24 | 85.82 |
| **1.1 – 3.0** | **155** | **122,877** | 0.29 | 96.76 |
| **3.1 – 5.0** | 54 | 34,832 | **0.34** | 51.16 |
| **> 5** | 19 | 9,324 | 0.33 | 25.47 |

\*Numbers in bold indicate where the highest values for a given factor occur, which imply increased priorities for vector control.

### Finding Optimized Delivery Routes

The most efficient routes of travel from mosquito control facilities to service demand locations were calculated using Dijkstra’s algorithm, modified to find the shortest routes between multiple origins (i.e. service facilities) and destinations (i.e. neighborhoods) as employed by the ‘Closest Facility’ tool in the ArcMap Network Analyst toolbox. Driving distance (km) along the road network was specified as the as the impedance, or cost of access to be minimized. Locations of individual households were not discernable from the aggregated data provided by INEC, and service destinations were set to the centroid of each neighborhood. Many of the block centroids did not intersect directly with the road network, and a 500 m tolerance was set to ensure the inclusion of all destinations in the analysis.

The monetary costs associated with delivery of mosquito abatement services in Machala were estimated for the optimized driving routes found in the Closest Facility analysis. In Machala, MoH mosquito control staff are capable of treating approximately 25 households with backpack sprayers before returning to a spray hub to refill, assuming one pair of spray technicians per deployment. Using this estimate of service capacity, the number of trips that MoH service teams need to make in order to completely treat every household in a given neighborhood was calculated by dividing the total number of households in a neighborhood by the number of houses treated in a single trip. Overall cost of access for neighborhoods was estimated by applying estimates of fuel consumption for service team trucks to the distance of optimized service delivery routes, multiplied by the number of trips needed to treat all households within a given neighborhood. Fuel economy was estimated using the average price of gasoline in Ecuador ($0.61/L in 2016) and the fuel consumption of a standard pickup truck manufactured in 2010 (5.53 km/L in city), values that reasonably reflect the price of gasoline and grade of service vehicles currently available to mosquito control teams in Machala [35,36].

#### Alternate Service Locations

Currently, mosquito control services are delivered from two hubs located in central Machala. To test if other combinations of locations may enable more efficient delivery of services under difference management strategies, we used the “Location-Allocation” tool in ArcGIS Network Analyst Toolbox. Location-allocation problems, where the best sites for service deployment are identified from a set of candidate locations, can be solved to meet a variety of user-specified goals, such as minimizing driving distance or maximizing the number of households served. We set the Location-Allocation tool to minimize weighted impedance, defined as driving distance along the road network, from deployment facilities to demand points (i.e. neighborhoods) with the goal of finding optimal placement for two spray hubs in Machala under different mosquito control strategies. Eight medical subcenters in Machala operated by the MoH were designated as candidates for alternate spray hub locations (Fig. 1). These subcenters were formerly outfitted for mosquito control operations prior to consolidation of abatement activities in Machala, making them logistically feasible for new potential locations of service deployment. Four location-allocation analyses were performed, where 1) only distance travelled on the road network was set as impedance without an additional weighting factor, identifying the two best locations to reduce overall transportation costs; 2) demand points (i.e. neighborhoods) were weighted by population size, identifying optimal locations to not only reduce driving distances, but also to prioritize those locations with highest demand; 3) demand points were weighted by reported human dengue cases, targeting areas that are prioritized for treatment under reactionary vector control; and 4) weighting demand by HCI, a scenario which simulates proactive management decisions based on a known social-ecological risk factor for dengue. The optimal facilities identified from these four location-allocation analyses were compared to current facility locations in terms of the relative cost and accessibility.

## Results

Mapping of census and epidemiological data revealed marked differences in the spatial distribution of factors that may be used to influence mosquito control decisions in Machala (Fig. 2). Indicators related to human population and settlement appear to be heterogeneous throughout the city, while the highest observations of dengue incidence were more centrally located during the 2010 outbreak. In contrast with reported dengue, households with high HCI (i.e. poor condition) are more peripherally located in Machala.

### Establishment of Service Areas

Given the centralized location of the two active mosquito abatement deployment facilities in Machala, estimated catchment areas of service based on driving distance from facilities radiate from the city’s center, indicating greater impedance to access of peripheral neighborhoods, in particular the Puerto Bolivar port region in the west (Fig. 3). The area within 1.1–3.0 km driving distance of either deployment facility encompasses the highest population (n=122,877), while the lowest population (n=9,324) was found more than 5 km driving distance from deployment hubs (Table 1). Neighborhoods with the highest quality housing (mean HCI=0.21) were located in central Machala, within 0.0–0.5 km driving distance of the spray hubs, while the poorest housing conditions (mean HCI=0.34) were found within 3.1 –5.0 km driving distance, in the urban periphery (Table 1).

**Fig. 3.**
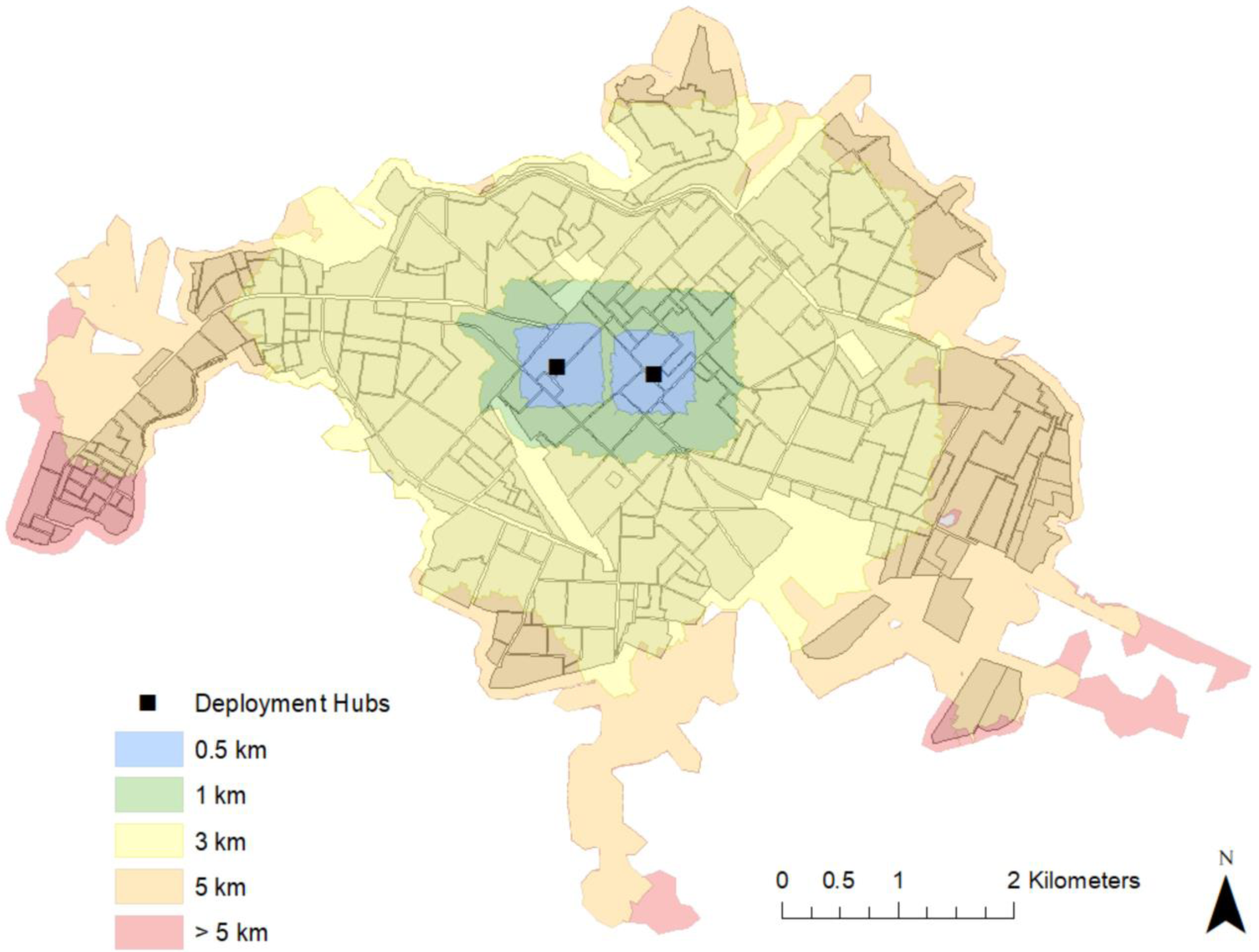
Service areas based on driving distance from the Ecuadorian Ministry of Health’s two centrally located spray deployment hubs in Machala, Ecuador. Each polygon represents the service catchment area associated with the corresponding driving distance along Machala’s road network. This figure was produced with modeled service area output in ArcMap 10.4 (ESRI, Redlands, CA).

### Finding Optimized Delivery Routes

The length of unidirectional spray routes, optimized to reduce distance, ranged from 0.14 km for neighborhoods near service deployment facilities, to 5.78 km for destinations near Machala’s municipal limits (Fig. 4).

**Fig. 4.**
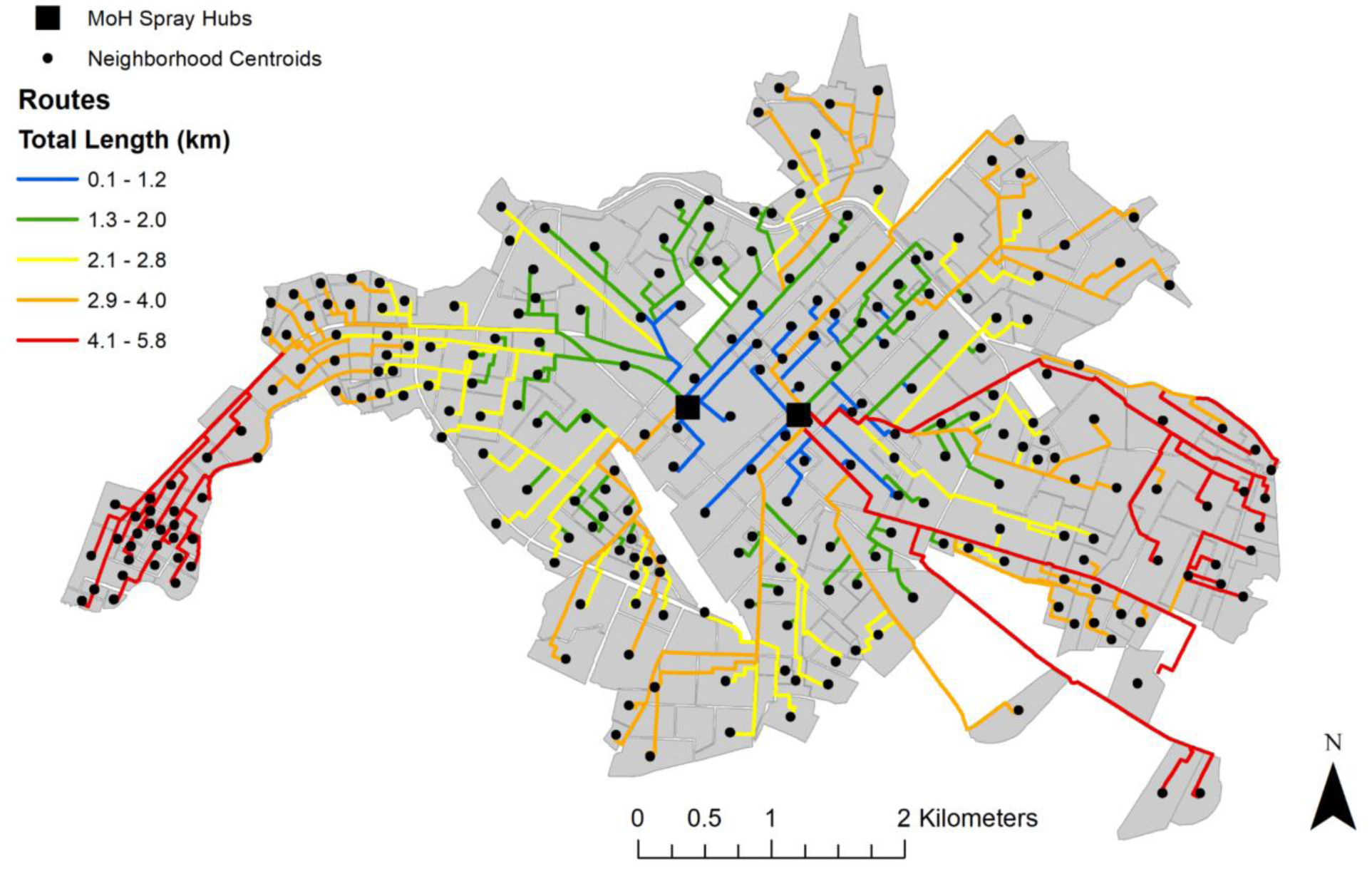
Optimized routes from closest spray hub in Machala based on driving distance, where the centroids of census blocks were specified as service destinations. This figure was produced with modeled route optimization output in ArcMap 10.4 (ESRI, Redlands, CA).

The centralized location of deployment facilities translates into generally increased driving distance, or impedance to access, for neighborhoods moving away from the city’s center. Applying fuel efficiency estimates to these distances, the cost associated traveling along optimal routes ranged from $0.02–$1.28 (USD), indicating the cost of gasoline consumed in one round trip to a given neighborhood. Applying fuel consumption estimates for optimized routes to the number of deployment trips needed to fully treat a neighborhood (i.e. where spray teams treat each household in a neighborhood once, returning to a hub to refill spray packs after treating 25 houses), enabled us to map and visualize the relative cost of accessing neighborhoods for treatment in the context of service demand (Fig. 5).

**Fig. 5.**
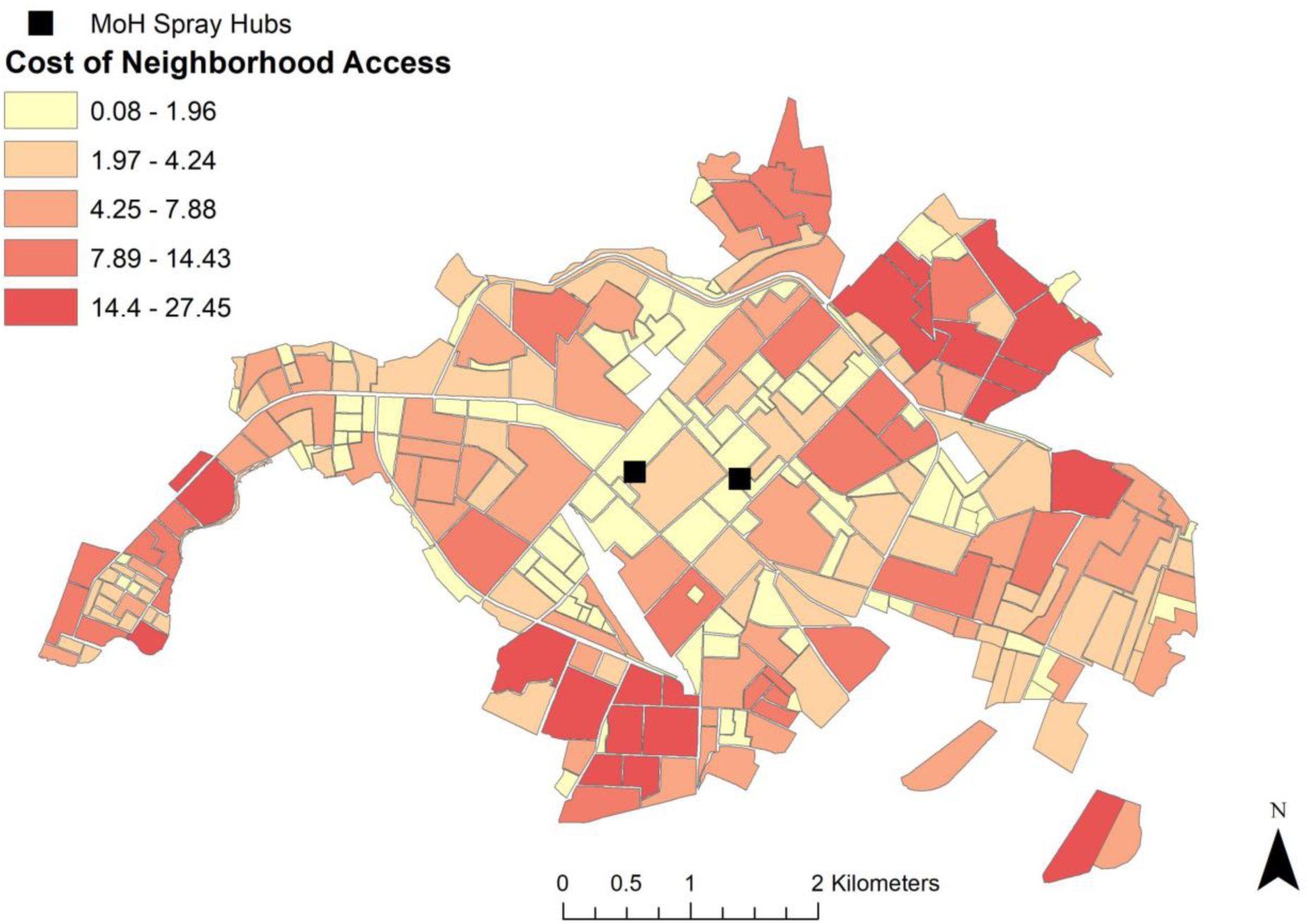
Estimated cost of service access for optimized driving routes from the closest mosquito spraying facility to neighborhood block centroids in Machala. Relative cost of access was determined via fuel consumption along routes and the number of trips required by mosquito control operators to treat each household in a neighborhood once, providing a visual means of comparing cost of access to demand for service. This figure was produced in ArcMap 10.4 (ESRI, Redlands, CA).

Estimated cost of access to treat all households in a given neighborhood block ranged from $0.08-$27.45, with an average cost of $4.03. Neighborhoods with the lowest cost of access had the highest dengue incidence, while neighborhoods with mid-range cost of access require the greatest number of deployments to treat all households (Table 2). However, remote neighborhoods with high cost access routes had the highest mean HCI, signifying the poorest quality housing (Table 2).

**Table 2.** Distribution of population, mean HCI, and 2010 dengue incidence for mosquito control service areas by optimized spray route costs.

| Route Cost<br>(\$ USD) | Neighborhoods | Population | Mean<br>HCI | Dengue Incidence<br>(per 10,000) |
| --- | --- | --- | --- | --- |
| **0.08 – 1.96** | **100** | 32,511 | 0.26 | **98.46** |
| **1.97 – 4.24** | 71 | 41,761 | 0.30 | 63.65 |
| **4.25 – 7.88** | 50 | **56,203** | 0.31 | 68.63 |
| **7.89 – 14.43** | 22 | 40,465 | 0.33 | 56.53 |
| **14.44 – 27.45** | 11 | 30,260 | **0.35** | 40.01 |

\*Numbers in bold indicate where the highest values for a given factor occur, which imply increased priorities for vector control.

### Alternate Service Locations

Location-allocation models demonstrate that the optimal combination of locations for mosquito abatement facilities changes, dependent upon specified management goals. When the goal was set to minimize the distance traveled along the road network, the current easternmost central hub is retained, while the western portion of the city is better serviced when control services are deployed from the subcenter located to the west of the currently active facility (Fig. 6A).

**Fig. 6.**
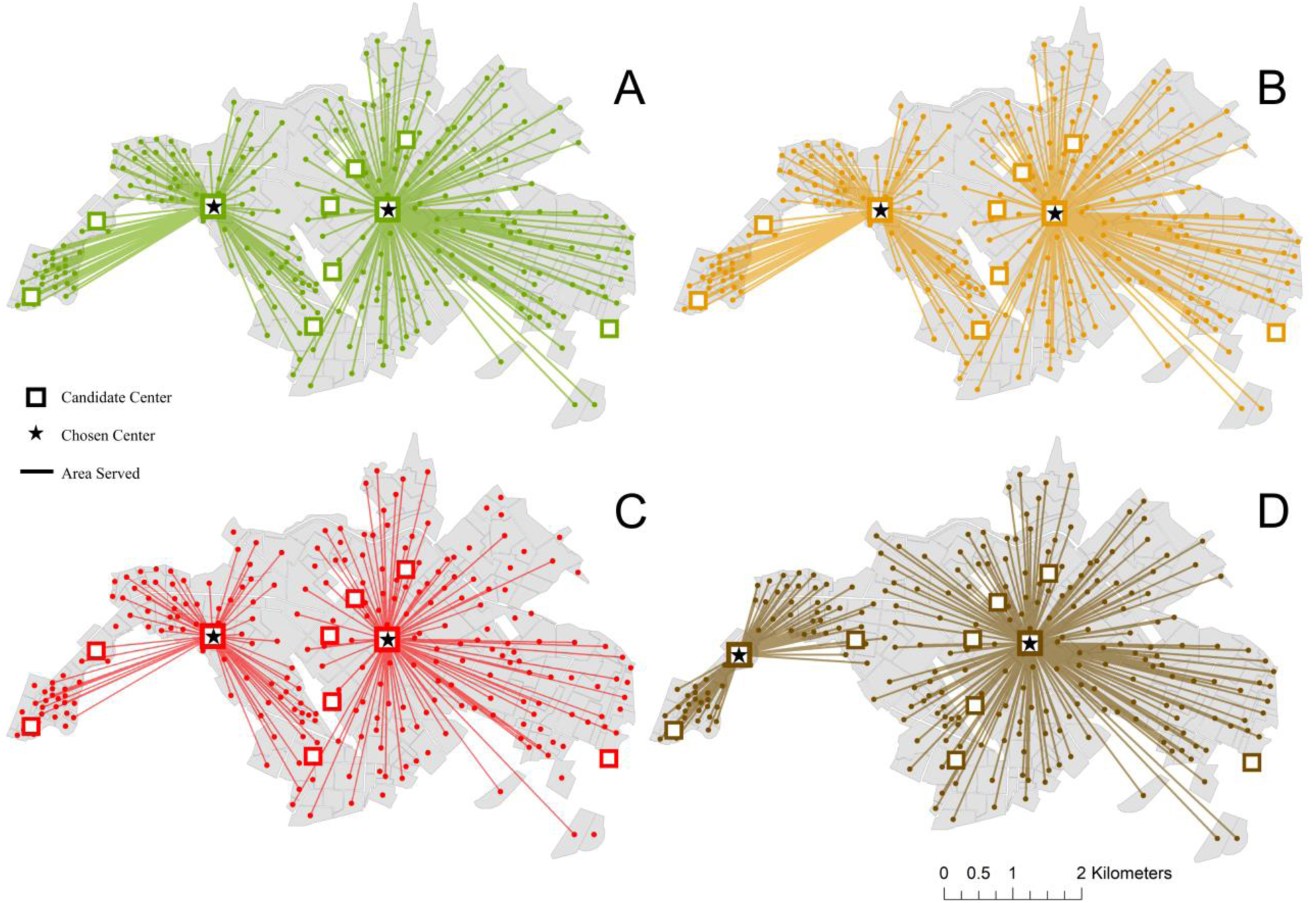
Location-allocation analysis results for Machala, where the best combination of facilities is chosen to minimize driving distance along the road network, prioritizing neighborhoods by distance to service hub (A), neighborhood population (B), dengue incidence (C), and housing condition (D). In each instance, one of the currently used locations is retained, while the second location for optimal delivery of mosquito abatement services depends on specified management priorities. This figure was produced with modeled optimized service locations in ArcMap 10.4 (ESRI, Redlands, CA).

These same locations were also selected as optimal for reducing driving distances when weighted by the population in each neighborhood, representing a management goal of reducing driving distances while prioritizing high-population areas (Fig. 6B), and when weighted by reported dengue incidence (Fig. 6C), representing surveillance-based reactive management. When weighting facilities by HCI, a scenario which represents a proactive mosquito control strategy, the easternmost current hub is again selected, while a subcenter even further to the west was selected as the complimentary location to optimize the tradeoff between distance and targeted housing conditions (Fig. 6D). Running a closest facility analysis for optimized routes on alternative spray hub locations weighted by distance, population, and dengue incidence, we found that estimated costs of fuel consumed on routes ranged from $0.02 to $1.06 per round trip, resulting in costs of accessing neighborhoods for treatment that ranged from $0.12 to $27.45, with an average cost of $3.52. These estimated costs of delivery from alternative hub locations indicate a 12.7% reduction in average fuel costs when compared to the currently active spray facilities, also resulting in a 33.7% increase in the population served by the least expensive routes (Table 3).

**Table 3.** Distribution of population, mean HCI, and 2010 dengue incidence for mosquito control service areas, based on estimated fuel consumption along optimized spray routes from alternative service locations found with Location-Allocation models.

| Location-Allocation<br>Weighting Factor | Route Cost<br>(\$ USD) | Neighborhoods | Population | Mean<br>HCI | Dengue<br>Incidence<br>(per 10,000) |
| --- | --- | --- | --- | --- | --- |
| Distance<br>Population<br>Dengue Incidence | 0.08 – 1.96 | 122 | 43,465 | 0.29 | 88.85 |
|  | 1.97 – 4.24 | 61 | 40,024 | 0.29 | 66.92 |
|  | 4.25 – 7.88 | 45 | 53,960 | 0.30 | 65.90 |
|  | 7.89 – 14.43 | 18 | 38,605 | 0.32 | 65.01 |
|  | 14.44 – 27.45 | 8 | 25,172 | 0.34 | 50.43 |
| Housing Condition | 0.08 – 1.96 | 125 | 43,857 | 0.29 | 87.25 |
|  | 1.97 – 4.24 | 56 | 38,052 | 0.29 | 47.82 |
|  | 4.25 – 7.88 | 45 | 50,568 | 0.30 | 83.57 |
|  | 7.89 – 14.43 | 19 | 40,076 | 0.30 | 85.58 |
|  | 14.44 – 27.45 | 9 | 28,647 | 0.34 | 61.14 |

Choosing facilities that optimized coverage of neighborhoods based on HCI, estimated fuel consumption for optimal routes ranged from $0.04 to $1.06 per round trip, resulting in costs of accessing neighborhoods for treatment that ranged from $0.12 to $27.45, with an average cost of $3.66. This estimated cost of delivery indicates a 9.2% reduction in average fuel costs compared to the currently active spray facilities, and a 34.9% increase in the population served by the least expensive routes (Table 3). Selected candidate locations not only lower the average costs associated with current optimized routes, but also lower the relative cost of access in the western urban periphery (Fig. 7).

**Fig. 7.**
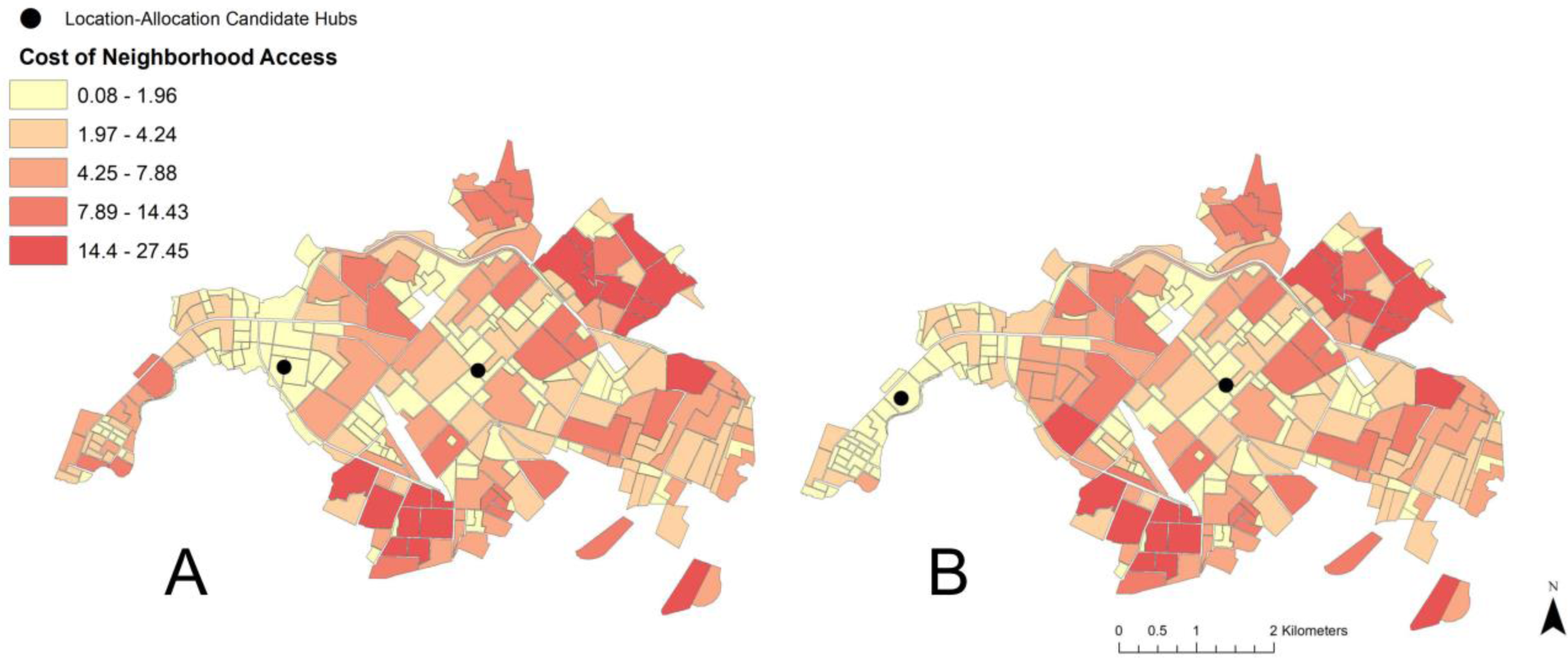
Estimated cost of service access for routes optimized under different candidate deployment locations in Machala, as determined via location-allocation analyses. Relative cost of access was determined via fuel consumption along routes and the number of trips required by mosquito control operators to treat each household in a neighborhood once. This figure was produced in ArcMap 10.4 (ESRI, Redlands, CA).

## Discussion

The results of this study highlight the importance and practical application of transportation network analysis to optimize mosquito control interventions in a dengue-endemic region. In visualizing census and epidemiological data from Machala by neighborhood blocks, there are clear discrepancies in the spatial distribution of factors commonly used by ministry officials to guide vector control decisions, particularly in poor quality housing, which is concentrated near the city’s outer limits (Fig. 2). The variation in spatial distributions of factors translates into differential prioritization of locations for treatment throughout the city, dependent upon specified management goals. Summarizing georeferenced data by transportation network service areas provided a general assessment of accessibility from spray deployment hubs, and demonstrated underlying differences that can impact decision-making and risk perception (Table 1, Fig. 3). For example, if the agency’s goal is to maximize the number of people or households that receive control services, then the service area within 1.1–3 km driving distance of active spray hubs would represent a high management priority. In contrast, if the abatement goal is to target households in poor condition, a strategy to disrupt transmission pathways via reduced exposure to vectors, then the service area within 3.1–5 km of spray hubs would be a more suitable target for concentrating treatments. Under the optimized route model for closest facilities, neighborhoods with the poorest HCI, and some areas of high population in northern and southern Machala, are among the costliest locations to access from current spray deployment hubs based on driving distance (Figs. 3 & 4). Cost of access in Machala, whether expressed as distance along the road network or estimated fuel consumption, is generally higher for the urban periphery, particularly near municipal boundaries (Figs. 4 & 5).

Visualizing census and epidemiological surveillance data in Machala reveals stark differences in the spatial distribution of reported dengue rates in the year 2010 and HCI, a known driver of arbovirus transmission in Machala (Fig. 2B & C). In a decision-making capacity, these factors represent two very different management philosophies in mosquito control—proactive versus reactive management. Mosquito abatement services are currently initiated at the beginning of coastal Ecuador’s rainy season in anticipation of mosquito production resulting from impounded water; individual neighborhoods are targeted in response to incoming human cases and areas of historically high risk. The MoH detects cases via passive arbovirus surveillance and not in a systematic fashion, and budgetary constraints on vector control and surveillance have become more pronounced following the dissolution of Ecuador’s national vector control program [10]. The MoH schedules regular larviciding in Machala. However, focal control in spatially discrete areas is very much reactive in nature, a response to cases detected via surveillance. Larvicides and adulticides are applied in critical locations only after human cases have been reported to clinics and verified by the MoH, well after transmission events have taken place. While this management strategy may aid in suppressing localized outbreaks and minimizing upfront abatement costs, it is nevertheless susceptible to inherent lags in surveillance systems and underreporting of cases, which are often exacerbated in at-risk areas with limited access to health services or low health care seeking behavior [37,38]. Indeed, the dearth of reported surveillance data in Machala’s periphery is counterintuitive, given that neighborhoods near the city’s municipal limits generally have elevated risk of exposure as determined via poor housing condition [39]. In exploring alternative management strategies, proactive mosquito abatement may be a viable addition to current policy, where neighborhoods with known risk factors are targeted for treatment before peaks in seasonal outbreaks are expected. However, under the current mosquito abatement structure, proactive management may not be the most cost-effective policy, as neighborhoods with the poorest quality housing have the largest economic barriers to access (Figs. 4 & 5).

This novel modeling effort has enabled us to make a first assessment of the costs associated with the delivery of mosquito spray services in Machala, using distance and fuel estimates as the impedance, or cost, of access. While this is a reasonable proxy of assessing relative barriers to access, several costs incurred by mosquito controllers were not included in our models due to lack of data, resulting in an underestimation of true operating expenses. Costs associated with abatement methods, driving times, time needed to treat each neighborhood, number of deployed technicians, agency operating hours, and quality of services could not be accounted for in these models. Data on the recurring costs associated with maintaining an operational fleet of vehicles (e.g. upkeep and repair costs) were also not available for this study [40]. In reality, the cost of access may be higher than estimated for some neighborhoods, particularly in the urban periphery where factors such as lack of paved roads may increase the time and resources needed to treat an area, as well as wear on fleet vehicles. We also assumed in these analyses that both spray hubs are fully operational, offering equivalent services. This is not always the case, particularly with delivery of ULV fogging services, as the number of operational vehicles in the spray fleet fluctuates due to mechanical issues. Furthermore, census and epidemiological data were only available for the most recent census year, which does not reflect current conditions, possible spatiotemporal shifts in disease risk, or potentially vulnerable communities residing beyond the official administrative limits of Machala [22,41]. The available spatial resolution of neighborhoods presents a further limitation in using these models for optimizing real-world service routes. Although not available for this study, georeferenced data on household locations within each neighborhood would allow us to better estimate the costs and driving distances associated with delivering household-level services, providing mapped routes that could conceivably be shared with mosquito control personnel.

Mapping optimal driving routes not only provides a means of streamlining service delivery, but also enables us to identify where high impedance to remote locations occurs. Excessive transportation distances may indicate practical barriers to service, limiting the number of people or vulnerable households that are able to effectively receive mosquito control services. This has implications for resource allocation and goal setting, where budgetary caps and personnel availability place logistical constraints on how often neighborhoods are treated. In Machala, delivery of abatement services in neighborhoods with poor housing condition, a previously described driver of mosquito production, becomes more costly, particularly for densely populated neighborhoods that require multiple visits to restock insecticides (Fig. 5) [16,25,42]. Previous studies conducted in Machala indicate that the urban periphery is not only more likely to have characteristics that drive dengue transmission, but also residents of these areas feel neglected with regards to mosquito control services offered by the MoH [10,42]. Accordingly, the centralized location of current mosquito abatement facilities is not ideal, in the sense that the shortest network distances overlap in areas that may not be high priority targets for treatment. While the best candidate locations for spray facilities are dependent upon desired impacts, results of the location-allocation analysis support that the current combination of mosquito abatement hubs is not selected as the most efficient in any of the tested management priorities. Still, there may be logistical benefits to the current location of facilities, such as personnel coordination, communication, and resource sharing. Therefore, we suggest that when selecting optimal locations for mosquito control facilities in the future, clear management goals and priorities must be defined for abatement programs.

## Conclusions

We have designed the first optimized transportation network for the delivery and assessment of mosquito control services in southern coastal Ecuador. The distance-based approaches used in this study, including formation of service areas, optimization of service routes, and exploration of goal-oriented management strategies, can serve as a template for other regions burdened with mosquito-borne disease transmission. The network analysis framework featured in this study highlights the utility of applying public health planning methodologies specifically to planning vector control programs. Resulting route optimizations and visualizations offer a powerful means of informing agency decision making, allowing for public health officials to critically assess the costs associated with delivery of services throughout a municipality. Additionally, the tools presented here offer a flexible environment in which current management strategies can be reviewed and compared to alternative policy approaches, systematically exploring possibilities for reducing network costs and improving accessibility in the face of limited agency resources.

## Data Availability

The data that support the findings of this study are available from INEC and the MoH, Ecuador, but restrictions apply to the availability of these data, which were used under license for the current study, and so are not publicly available. Data are however available from the authors upon reasonable request and with permission of INEC and the MoH, Ecuador.

## List of Abbreviations

YFV: yellow fever virus
DENV: dengue virus
CHIKV: chikungunya virus
ZIKV: Zika virus
SNEM: Servicio Nacional de Control de Enfermedades Transmitidas por Vectores Artrópodos
MoH: Ministry of Health
ULV: ultra=low volume
INEC: Instituto Nacional de Estadística y Censos
HCI: housing condition index

## Declarations

### Ethics approval and consent to participate

This protocol was reviewed and approved by Institutional Review Boards (IRBs) at SUNY Upstate Medical University, the Luis Vernaza Hospital in Guayaquil, Ecuador, and the MOH of Ecuador. Prior to the start of the study, all participants engaged in a written informed consent or assent process, as applicable. IRB number: [417710] Capacity Strengthening in Ecuador: Partnering to improve surveillance of febrile vector-borne diseases.

### Consent for publication

Not Applicable

### Competing interests

The authors have no competing interests.

## Funding

CAL was supported by NSF DEB EEID 1518681. The funders had no role in study design, data collection and analysis, decision to publish, or preparation of the manuscript.

## Authors’ contributions

CAL, LM, and SJR conceived of the study. CAL, NH, AMS, and EBA compiled the data used in analyses. CAL conducted analyses and drafted the manuscript. CAL, LM, NH, AMS, NDBC, JKB, and SJR assisted with interpretation of the data and provided feedback for this manuscript. All authors read and approved the final manuscript.

## Acknowledgements

Many thanks T. Ordoñez and staff at the Ministry of Health in Machala, Ecuador, who provided data for this study.

## Notes

### Competing Interest Statement

The authors have declared no competing interest.

### Author Declarations

All relevant ethical guidelines have been followed and any necessary IRB and/or ethics committee approvals have been obtained.

Any clinical trials involved have been registered with an ICMJE-approved registry such as ClinicalTrials.gov and the trial ID is included in the manuscript.

